# Intraocular water movement visualization using ^1^H-MRI with eye drops of O-17 labeled saline: a first-in-human study

**DOI:** 10.1101/2022.02.13.22270420

**Authors:** Moyoko Tomiyasu, Yasuka Sahara, Etsuko Mitsui, Hiroki Tsuchiya, Takamasa Maeda, Nobuhiro Tomoyori, Makoto Kawashima, Toshifumi Nogawa, Riwa Kishimoto, Yuhei Takado, Tatsuya Higashi, Atsushi Mizota, Kohsuke Kudo, Takayuki Obata

## Abstract

**Background:** Visualization of aqueous humor flow in the human eye is difficult because gadolinium, a magnetic resonance imaging (MRI) contrast agent, does not readily cross the blood-retinal and blood-aqueous barriers of capillaries that supply water to the eye. The proton (^1^H) of oxygen-17 water (H_2_^17^O) has a very short transverse relaxation time (T_2_), and the T2-weighted (T2W) ^1^H-MRI signal intensity of a region with H_2_^17^O is lower than that with only H_2_ ^16^O.

**Purpose:** To observe the distribution of H_2_^17^O in the human eye, and the flow into and out of the anterior chamber of H_2_^17^O, using dynamic T2W ^1^H-MRI.

**Materials and Methods:** Seven ophthalmologically normal adult volunteers under 40 years old participated in this study. During dynamic imaging, the subject self-administered 10 mol% H_2_^17^O saline (0.92–1.37 mL) to their right eye for 1 min. Time-series images were created by subtracting the image before the eye drops from each of the images obtained after the eye drops. The “normalized signal intensity of the right anterior chamber” (rAC) in each image was obtained by dividing the signal intensity of the right anterior chamber region-of-interest by that of the left. Changes in transverse relaxation rate and H_2_^17^O concentration (*P*_*O*17_) were calculated from the rAC. The inflow and outflow constants of H_2_^17^O in the right anterior chamber were also determined.

**Results:** Decreased signal intensity after the H_2_^17^O eye drops was observed in the anterior and posterior chambers, but not in the vitreous body. The rAC signal intensity decreased after the eye drops, and then recovered to close to rAC(0) at 40 min. The inflow and outflow constants were 0.53 ± 0.19 and 0.055 ± 0.019 min^-1^, respectively, and the peak value of *P*_*O*17_ was 0.19 ± 0.04% (mean ± SD).

**Conclusion:** H_2_^17^O saline eye drops distributed in the human anterior and posterior chambers. Further, the eye drops smoothly flowed into, and slowly out of, the anterior chamber.

**Summary:** O-17 water dropped into human eyes as a T2-weighted MRI contrast agent was distributed in the anterior and posterior chambers, with smooth flow into, and slow outflow from, the anterior chamber.

**Key findings:** In seven healthy volunteers, dynamic T2-weighted ^1^H-MRI showed that the signal intensity in the anterior chamber decreased smoothly after H_2_^17^O eye drops and then slowly recovered, reaching a value close to that observed before the eye drops, after 40 minutes, with inflow and outflow constants of H_2_^17^O of 0.53 ± 0.19 min^-1^ and 0.055 ± 0.019 min^-1^, respectively. The signal changes were limited to the anterior and posterior chambers and were not observed in the vitreous body.

## Introduction

Abnormalities in the aqueous flow in the eye cause eye diseases such as glaucoma (1). If the flow of aqueous humor could be visualized, it would not only help in the diagnosis of glaucoma, but could also help with diagnoses and therapeutic effects for various other ocular diseases. Such visualization of water flow would also be expected to make a significant contribution to drug discovery.

Gadolinium, as a magnetic resonance imaging (MRI) contrast agent, does not pass smoothly through the blood-retinal barrier or the blood-aqueous barrier of the capillaries that supply water to the eye, and leakage of gadolinium from the ora serrata into the vitreous has been reported with aging (2). Therefore, although gadolinium can be used to assess deterioration of the barriers due to aging and other factors, it is difficult to use it to evaluate aqueous humor flow. Oxygen-15-labeled water used in positron emission tomography may be used to evaluate animal eyes, but it is not suitable for human eyes because of its short half-life (about 2 min). D_2_O saline solution has been administered to some animals *in vivo* and the dynamics of the aqueous humor evaluated (3, 4). However, it is unethical to apply D_2_O in humans because of its toxicity (5).

The protons in oxygen-17-labelled water (H_2_^17^O) have six Lamour frequencies due to the scalar coupling with^17^O, which has a spin quantum number of 5/2, and the T_2_ of the H_2_^17^O proton is very short (6-10). The chemical exchange of protons between H_2_^17^O and H_2_^16^O also shortens the T_2_ of the H_2_^16^O protons. As a result, the T2-weighted (T2W) proton MRI (^1^H-MRI) signal intensity of a region with H_2_^17^O is lower than that with only H_2_^16^O. Several *in vivo* experiments using dynamic T2W ^1^H-MRI (dT2W_MRI) of H_2_^17^O have been reported (11-13), and, recently, an *in vivo* experiment on the human central nervous system has also been reported (13).

We hypothesized that after H_2_^17^O eye drop, the region with H_2_^17^O in the eye would have low signal intensity on T2W images therefore the flow of H_2_^17^O could be traced by dynamic imaging. Accordingly, our study using H_2_^17^O saline eye drops was designed to observe the distribution of H_2_^17^O in the eye, and to calculate its inflow and outflow ratios in the anterior chamber using dT2W_MRI.

## Methods

### Subjects

Seven healthy ophthalmologically normal adult volunteers under 40 years old participated in this prospective study. Institutional Review Board approval and written informed consent were obtained.

### Hardware

All ^1^H-MRI was performed using a clinical 3-T MR scanner (123.23 MHz; Magnetom Verio; Siemens Healthineers, Erlangen, Germany). A whole-body coil (bore diameter: 70 cm) and a 32-channel head coil (inner diameter: 22 cm) were used for radio frequency signal transmission and reception, respectively.

### Dynamic T2W ^1^H-MRI

Prior to the dT2W_MRI for the eye, T1-weighted sagittal 3-dimensional (3D) scout images (repetition time (TR)/echo time (TE), 3.15/1.37 msec; field-of-view (FOV), 260×260 mm; slice thickness, 1.6 mm; and matrix, 160×160×128) were recorded. The parameters of the dT2W_MRI were as follows: a half-Fourier, single-shot, turbo-spin echo sequence (14), TR/TE, 3000/444 msec; FOV, 180×180 mm; slice thickness, 3 mm; and matrix, 320×320 pixels, reconstructed to 640×640. During the dT2W_MRI, the subjects applied 10 mol% H_2_^17^O saline drops (0.92–1.37 mL; Taiyo Nippon Sanso, Tokyo, Japan) to their right eye for 1 min. Before and after the eye drops, the subjects were required to stare at a single point, to avoid eye movements. However, of the 3-sec TR, the time required for image acquisition was about 1 sec, and the subject could blink during the rest of the time. As this was the first trial to monitor H_2_^17^O in the human eye using ^1^H-MRI, the dT2W_MRI imaging protocol was modified during the course of the study, resulting in 3 patterns of sequential actions by the subjects and total scan times of 12 or 42 min (Fig. 1). In the 2 protocols with the 42 min scan time, some periods were inserted in which the subjects could have a rest (Eyes_closed).

**Figure 1.**
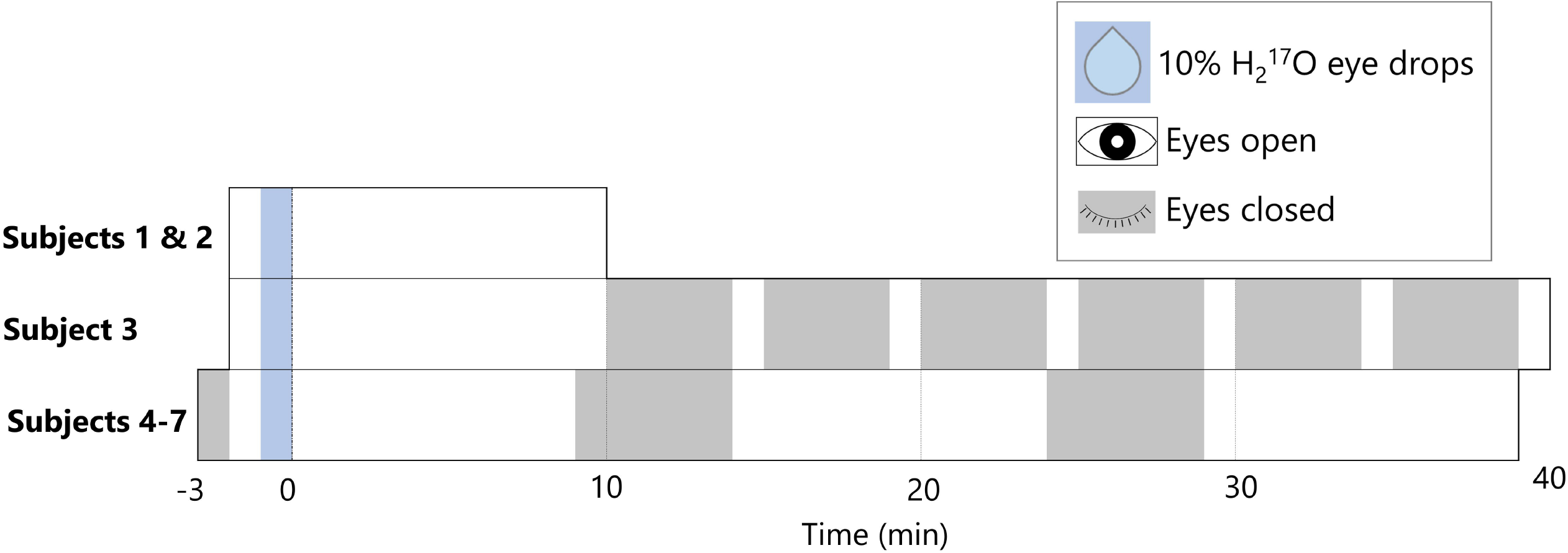
Sequences of actions for the subjects during dynamic T2-weighted ^1^H magnetic resonance imaging. Time t = 0 is the end of the application of H_2_^17^O eye drops.

### Signal quantification of the dynamic images

In-house software developed in MATLAB (MathWorks, Natic, MA) was used for the data analysis. First, the images were registered by the 2D affine transformation method (imregdemons in MATLAB). To investigate the variation across time in the signal intensities of the images, a time series of subtracted images were created as follows: each average image was created from 8–10 consecutive images obtained at a certain time; then the averaged image obtained prior to the eye drops was subtracted from each of those images. Three regions of interest (ROIs), located in the left and right anterior chambers and the left vitreous body, each of size 1.66 mm^2^ (7×3 pixels), were selected manually in the registered image (Supplementary Fig. 1). The normalized signal intensity of the right anterior chamber (rAC) for each image was obtained by dividing the signal intensity of the ROI of the right anterior chamber by that of the left. A linear relationship between H_2_^17^O concentration and the transverse–relaxation-rate change of the H_2_O proton (Δ*R*_2, *H*_), obtained from the MR signal intensity change, has been reported (7, 12). In our study, the Δ*R*_2, *H*_ of rAC was calculated using the following equation (12):

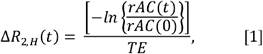

where rAC(0) is the signal intensity before the eye drops and TE = 0.444 sec is the dT2W_MRI parameter used in our study. The transverse relaxation rate of the H_2_^17^O proton 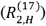 with 100% concentration was calculated using the following equation (9,15):

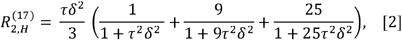

where τ is the proton-exchange lifetime, which is assumed to be 1.8 × 10^−3^ sec^−1^ (6); 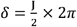, and J is the^17^O-^1^H scalar-coupling constant, which was measured as 91 Hz (16). The transverse relaxation of the H_2_O proton (*R*_2,*H*_) in the presence of H_2_^16^O and H_2_^17^O can be described by the following equation (15):

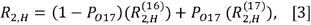

where *P*_*O*17_ is the molar fraction of H_2_^17^O, and 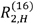 is the transverse relaxation rate of the H_2_^16^O proton. The Δ*R*_2, *H*_ at time = t is represented by the following equation:

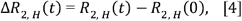

where *R*_2,*H*_(0) is the value at the time before the eye drops and is the same as 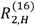. Using equations [1] to [4], and the fact that 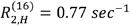 (17), *P*_*O*17_ in the right anterior chamber ROI was calculated. For the interval during which *P*_*O*17_ increased, the following exponential equation was fitted, to obtain the inflow constant of H_2_^17^O:

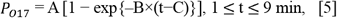

where A, B and C are constants; B is the inflow constant. A represents the maximum concentration of H_2_^17^O and C represents the gap time between the end of H_2_^17^O administration and the beginning of the *P*_*O*17_ change. Conversely, during the period when *P*_*O*17_ was decreasing, the outflow constant was obtained by fitting the following exponential equation:

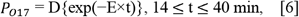

where D and E are constants; E is the outflow constant. Since the period from 9 or 10 min to 14 min was the Eyes_closed period, or the dT2W_MRI acquisition was finished at 10 min for some subjects, 1 ≤ t ≤ 9 min and ≥ 14 min were used to calculate the inflow and outflow constants, respectively (Fig. 1).

The quality criteria for each image data and series of data are as follows: the images obtained during the Eyes_closed periods, and the images with motion artifacts were not used in the analyses (Supplementary Figs. 1 and 2). Images with motion artifacts were those in which the signal intensity in the left vitreous ROI was within the outlier range defined by the interquartile range (IQR) method. The IQR was the difference in the values between 25% (Q1) and 75% (Q3) in the left vitreous ROI across all images, after excluding the images obtained during the Eyes_closed periods in each subject. Values smaller than Q1 − 1.5 × IQR or larger than Q3 + 1.5 × IQR were classified as outliers (Supplementary Fig. 1). The dT2W_MRI data with a coefficient of determination (R^2^) of less than 0.1 for the fitting curve of equation [5] or [6] were excluded.

## Results

dT2_MRI data were obtained from each of seven subjects, but the data that did not meet quality criteria were excluded, resulting in a five final data set. One of them had a short acquisition time of 12 min and was used only for the calculation of inflow constants.

Decreased signal intensity in the dT2W_MRI after the H_2_^17^O eye drops was observed in the right anterior and posterior chambers but not in the right vitreous body, as shown in Figure 2. The rAC signal intensity decreased after the eye drops, and then began to recover, reaching a value close to rAC(0) at 40 min (Fig. 3).

**Figure 2.**
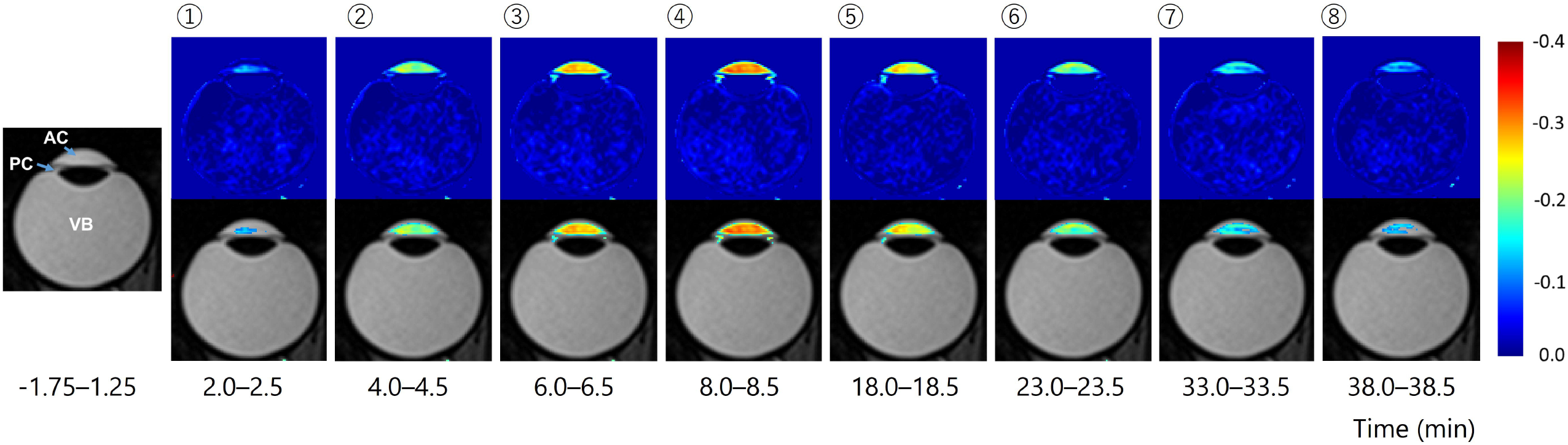
Representative T2-weighted (T2W) ^1^H magnetic resonance imaging (MRI) and sequential subtracted images from the right eye of a subject in their 20’s. <u>Left-most</u>: T2W image obtained before the application of the H_2_^17^O eye drops. <u>Upper row</u>: each image is the averaged image, obtained during the time period shown at the bottom of the column, subtracted from the averaged image obtained before the H_2_^17^O eye drops (left-most). Time t = 0 is the end of the eye drops. <u>Lower row</u>: each of the corresponding images in the upper row superimposed on a T2W image. The time axis of the averaged-image acquisition is shown in Figure 3. AC = anterior chamber, PC = posterior chamber, VB = vitreous body.

**Figure 3.**
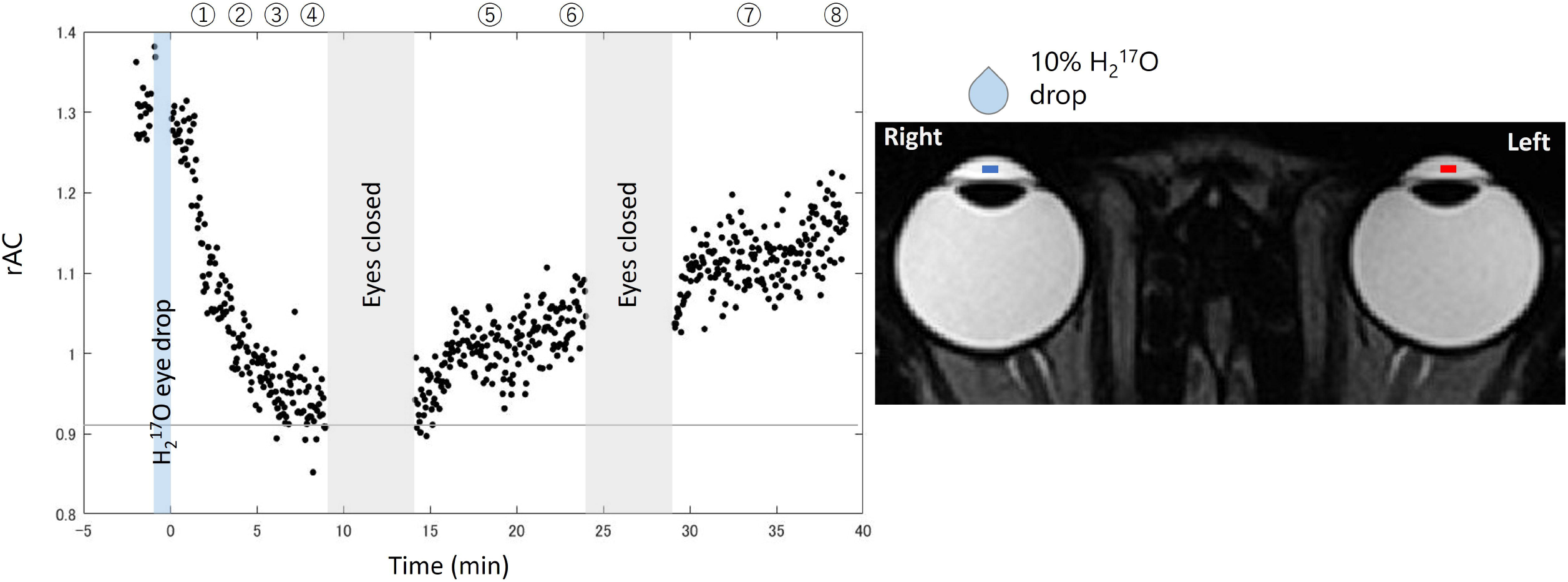
Representative variation across time in the normalized signal intensity in the right anterior chamber (rAC) obtained from dynamic T2-weighted ^1^H magnetic resonance imaging (a subject in their 20’s). The signal intensity was obtained by dividing the signal from the right anterior chamber region of interest (ROI) by that from the left. The blue and red areas in the upper-right image show the ROIs (1.66 mm^2^, each) for the right and left eyes, respectively. Time t = 0 is the end of the eye drops. The labels (➀ – ➇) at the top of the plot indicate the data-collection times for creating the subtracted images in Figure 2.

Combining the results from each subject, the time-variation values in H_2_^17^O concentration in the right anterior chamber were as follows [mean ± SD (range)]: inflow constant (n = 5), 0.53 ± 0.19 min^-1^ (0.37–0.87 min^-1^); outflow constant (n = 4), 0.055 ± 0.019 min^-1^ (0.038–0.072 min^-1^); maximum concentration (n = 5), 0.19% ± 0.04% (0.14%–0.24%) (Figs. 4 and 5).

**Figure 4.**
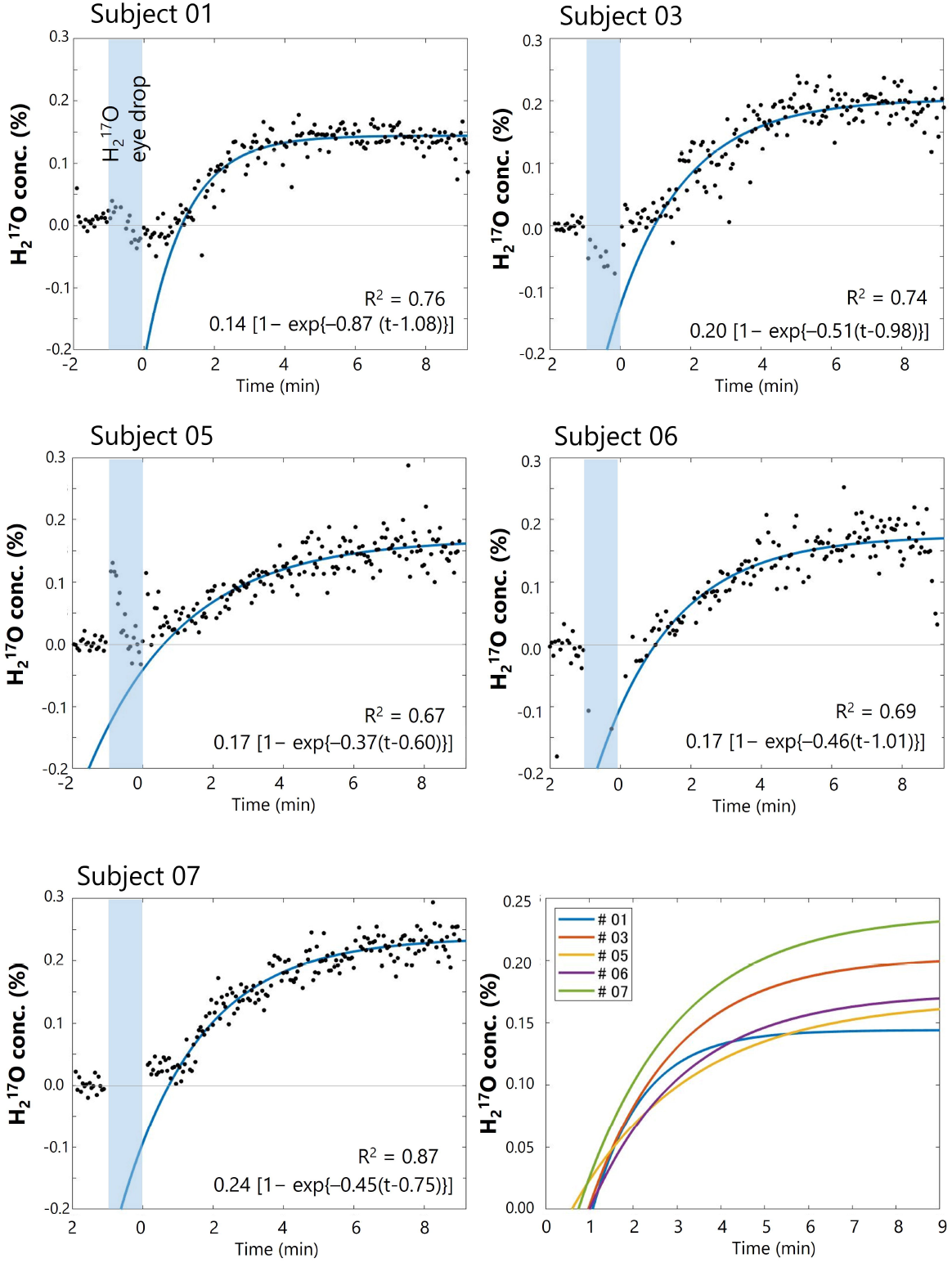
Time variations (−2 ≤ t ≤ 9 min) in the H_2_^17^O concentration [conc.(%)] of the right anterior chamber. Time t = 0 is the end of the eye drops. The exponential curves were fitted with equations of the form A [1 − exp{–B×(t−C)}], fitted at time (1 ≤ t ≤ 9 min), the parameters for which are shown on the graphs. R^2^ is the coefficient of determination. <u>Bottom-right:</u> plots of all fitting curves.

**Figure 5.**
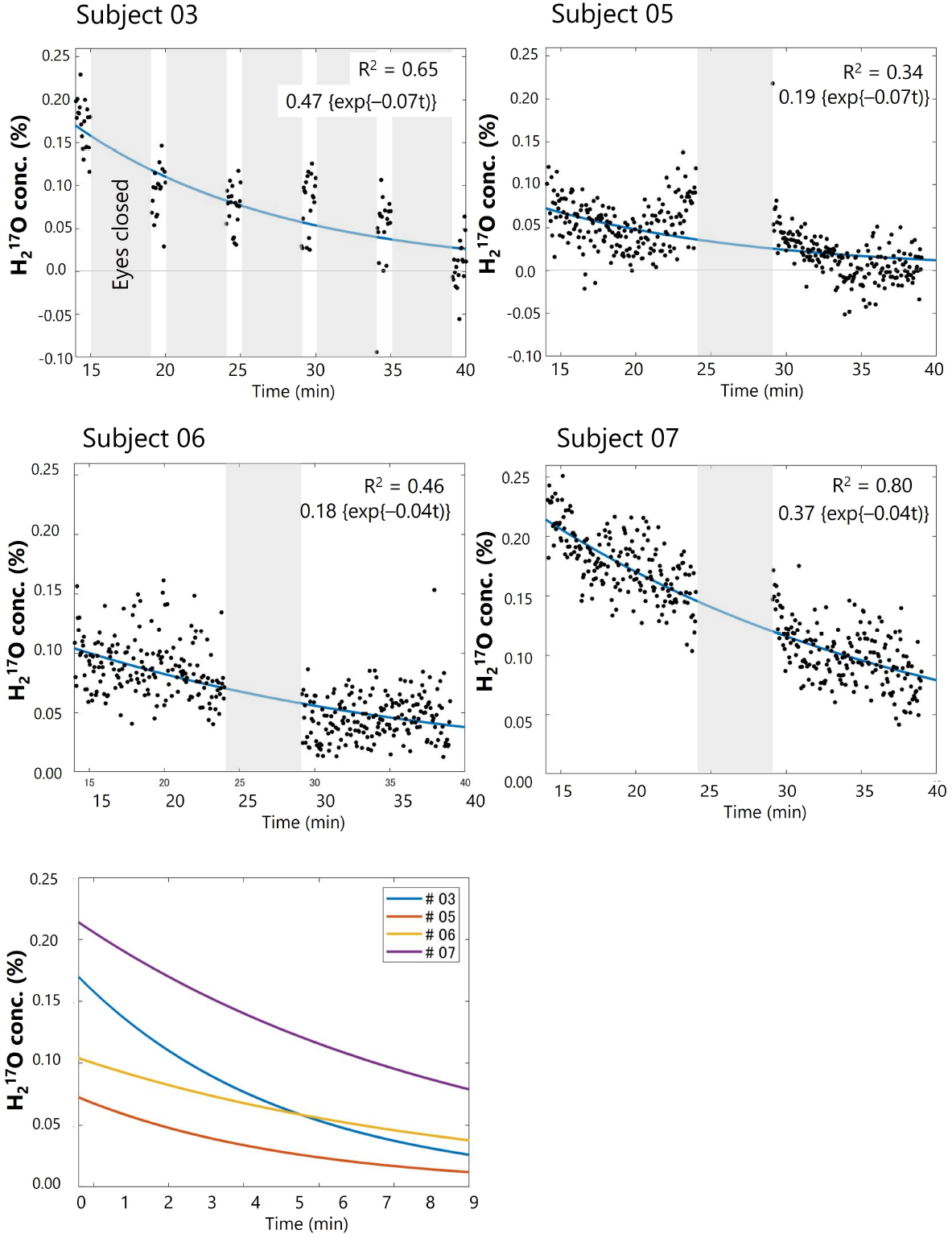
Time variations (14 ≤ t ≤ 40 min) in the H_2_^17^O concentration [conc.(%)] of the right anterior chamber. Time t = 0 is the end of the eye drops. The data were fitted with the exponential curve D{exp(−E×t)}, with the parameters shown on the plots. R^2^ is the coefficient of determination. <u>Bottom raw</u>: plots of all fitting curves.

## Discussion

In our prospective study, we quantitively assessed time-variation changes of H_2_^17^O concentration in *in vivo* human eye using dT2W_MRI, which was performed with a clinical 3-T scanner. There did not seem to be any change in the signal intensity in the left anterior chamber associated with the eye drops (Supplementary Fig. 1).

The ranges of the inflow and outflow constants were 0.37–0.87 min^-1^ and 0.02–0.07 min^-1^, respectively. Although there were substantial individual differences, the inflow constant was larger than the outflow constant in each subject as well as the averages, indicating that H_2_^17^O saline eye drops flow smoothly into the anterior chamber, and then flow slowly out again.

There have been no reports of H_2_^17^O being administered to human eyes *in vivo*, but there is one report in animal eyes (12). Kwong et al. administered H_2_^17^O eye drops to rabbits, and observed water movement in the rabbits’ eyes using 1.5-T MRI (12). The authors determined the H_2_^17^O outflow constants from the anterior chamber using both ^1^H-MRI and^17^O-MR spectroscopy, and found values of 0.084 and 0.107 min^-1^, respectively (12). Similar observations in rabbit eyes were reported by Obata et al.(4) in ^2^H-MRI with D_2_O. In their study, the outflow constant of D_2_O from the rabbit anterior chamber was 0.113 ± 0.017 min^-1^ (n = 4), which was similar to that with H_2_^17^O in Kwong et al.’s study (12). In our human study, the outflow constants [0.055 ± 0.019 min^-1^, 0.038–0.072 min^-1^, (n = 4)] were smaller than the earlier rabbit values, suggesting that water may flow out more slowly from the anterior chamber in humans than in rabbits. However, the coefficient of determination (R^2^) for the outflow constant was smaller than that of the inflow constant for each subject (Figs. 4, 5), suggesting that the outflow constant may also be affected by data instability resulting from the very long scan times.

Regarding the time variation of the dynamic ^1^H-MR images in our H_2_^17^O study, the signal changes were limited to the anterior and posterior chambers and were not observed in the vitreous body (Fig. 2). In the H_2_^17^O experiment in rabbit eyes, only the signal reduction and recovery in the anterior chamber by ^1^H-MRI was described (12). In a D_2_O study in rabbits, signal change in the vitreous body was observed, in addition to the changes in the anterior and posterior chambers (4). Considering those results, although the possibility that H_2_^17^O flowed into the vitreous in the present study cannot be denied, the signal changes would only have been at the noise level.

The method used in the present study also has the potential to evaluate the rate of aqueous humor exchange such as in diseases such as glaucoma, and in diseases during their treatment with drugs. However, it took about 40 min to monitor the H_2_^17^O saline washing out of the anterior chamber. When applying this method to a patient, it is necessary for them to keep their eyes opened and fixated for a long period, which is burdensome and may also result in increased motion artifacts. The development of less-burdensome imaging methods, including shorter imaging times, may make it possible to acquire more-stable MR images with fewer motion artifacts.

## Conclusion

Our results using dT2W_MRI showed that H_2_^17^O saline eye drops were distributed in the human anterior and posterior chambers. Also, the eye drops smoothly flowed into, and slowly out of, the anterior chamber. Further measurements on healthy volunteers are needed to confirm the reliability of the values obtained in this study.

## Data Availability

All data in the present study are available from the corresponding author upon reasonable request.

## Abbreviations

dT2W_MRI: dynamic T2W ^1^H-MRI
Δ*R*_2, *H*_: transverse relaxation rate change of the H_2_O proton
H_2_^17^O: oxygen-17 water
IQR: interquartile range
*P*_*O*17_: the molar fraction of H_2_^17^O
*R*_2, *H*_: transverse relaxation rate of the H_2_O proton
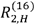: transverse relaxation rate of the H_2_ ^16^O proton
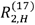: transverse relaxation rate of the H_2_^17^O proton
rAC: normalized signal intensity of the right anterior chamber
ROI: region of interest

## Acknowledgments

The authors thank Ms. S. Kawakami and Ms. H. Yamashita, Dr. H. Tsuji, and Dr. M. Makishima for their support of our clinical trial study. This work was supported by a Grant-in-Aid (Public/Private R&D Investment Strategic Expansion PrograM: PRISM) from the Cabinet Office, Japan. We thank Claire Barnes, PhD, from Edanz (https://jp.edanz.com/ac) for editing a draft of this manuscript.

## Figure Legends

**Supplementary Figure 1.**
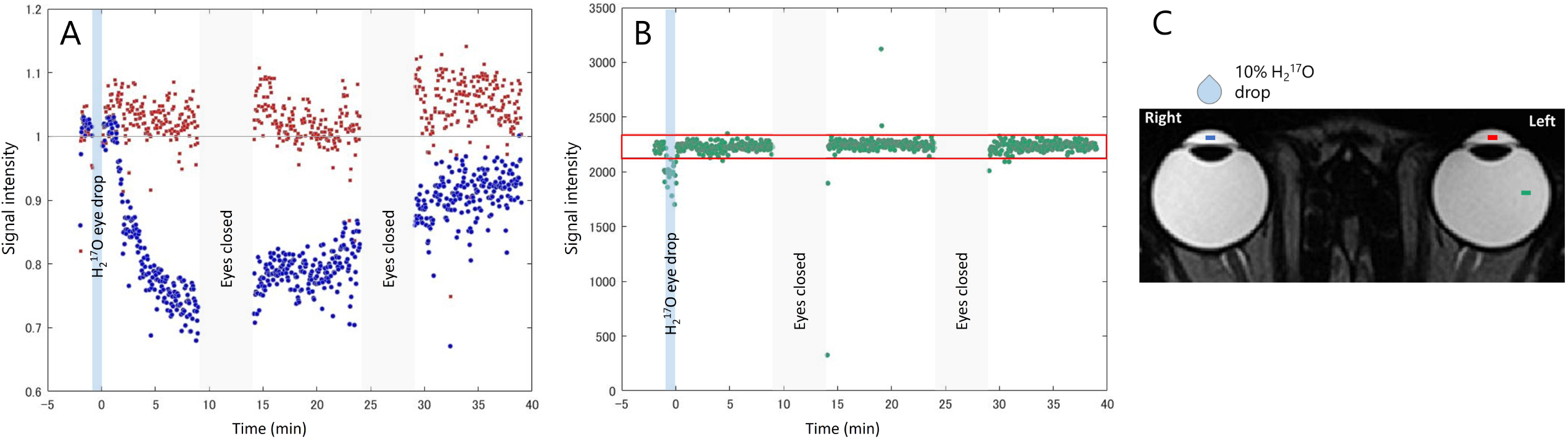
Representative time variations in the signal intensities of regions of interest (ROIs) of dynamic T2-weighted ^1^H magnetic resonance imaging (a subject in their 20’s). A: Signal intensities in the the right (blue) and left (red) anterior chambers. The signal intensities were normalized by dividing each by the signal intensity before the eye drops. B: Signal in the left vitreous body (green). The images with signal intensities outside the red frame are outliers in the interquartile range method, and were not used in the data analysis. C: the ROIs of each region are shown on the image (1.66 mm^2^, each).

**Supplementary Figure 2.**
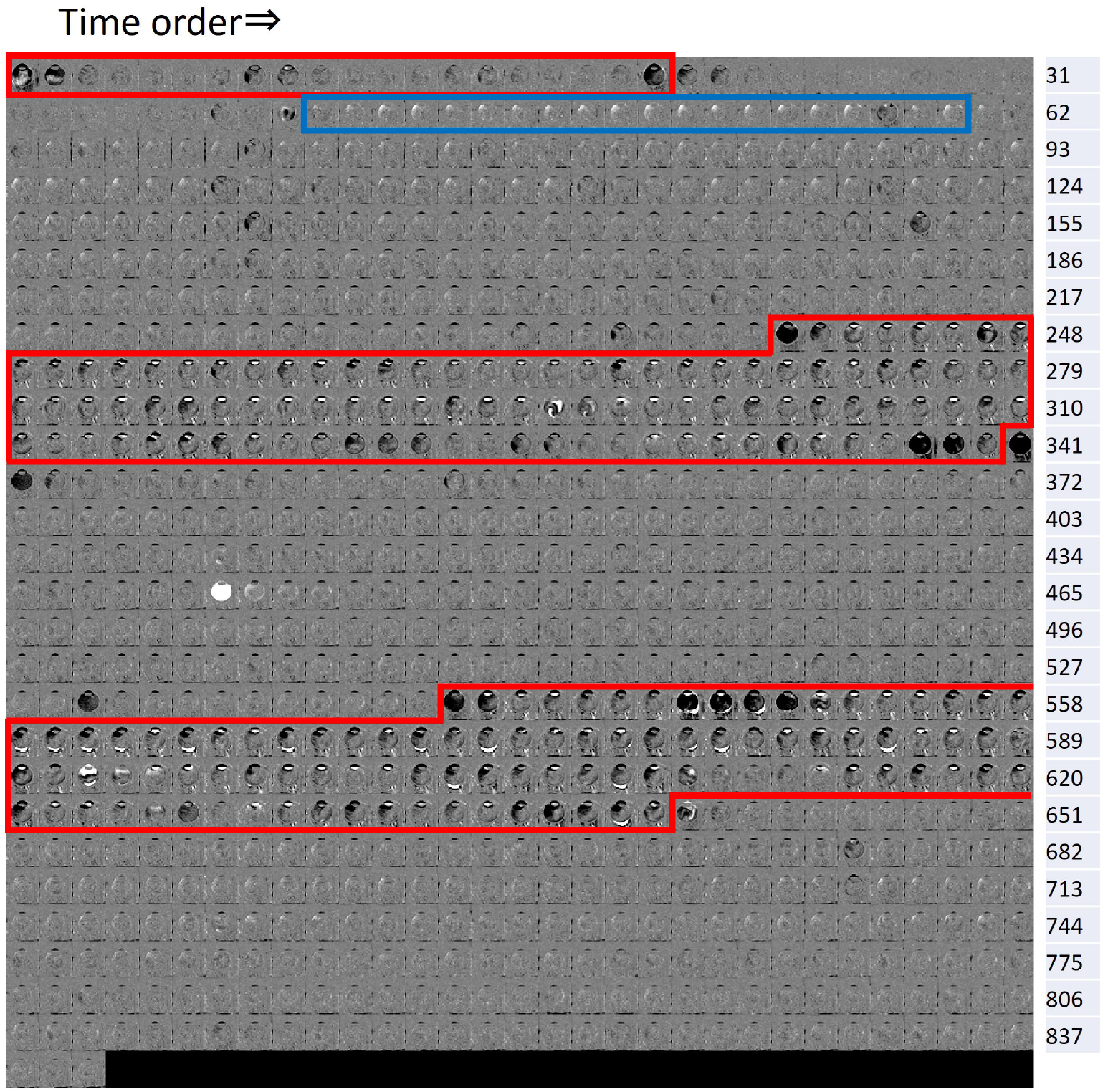
Representative time-series images of dynamic T2-weighted ^1^H magnetic resonance imaging (a subject in their 20’s). Each number on the right corresponds to the data-collection number of the images to the left. The images in the blue frame were obtained during the H_2_^17^O saline-drop period. The images in the red frames were rest periods (Eyes_closed), which were not used in the data analysis.

